# Dynamic Network Strategies for SARS-CoV-2 Control on a Cruise Ship

**DOI:** 10.1101/2020.08.26.20182766

**Authors:** Samuel M. Jenness, Kathryn S. Willebrand, Amyn A. Malik, Benjamin A. Lopman, Saad B. Omer

## Abstract

SARS-CoV-2 outbreaks have occurred on several nautical vessels, driven by the high-density contact networks on these ships. Optimal strategies for prevention and control that account for realistic contact networks are needed. We developed a network-based transmission model for SARS-CoV-2 on the Diamond Princess outbreak to characterize transmission dynamics and to estimate the epidemiological impact of outbreak control and prevention measures. This model represented the dynamic multi-layer network structure of passenger-passenger, passenger-crew, and crew-crew contacts, both before and after the large-scale network lockdown imposed on the ship in response to the disease outbreak. Model scenarios evaluated variations in the timing of the network lockdown, reduction in contact intensity within the sub-networks, and diagnosis-based case isolation on outbreak prevention. We found that only extreme restrictions in contact patterns during network lockdown and idealistic clinical response scenarios could avert a major COVID-19 outbreak. Contact network changes associated with adequate outbreak prevention were the restriction of passengers to their cabins, with limited passenger-crew contacts. Clinical response strategies required for outbreak prevention included early mass screening with an ideal PCR test (100% sensitivity) and immediate case isolation upon diagnosis. Public health restrictions on optional leisure activities like these should be considered until longer-term effective solutions such as a COVID-19 vaccine become widely available.

## INTRODUCTION

COVID-19, caused by SARS-CoV-2, is responsible for large-scale morbidity and mortality across the world. In addition to virus spread from symptomatic individuals, asymptomatic and pre-symptomatic transmission has also been documented (1). At an individual level, living in close proximity to someone with COVID-19 increases risk of contracting the disease (2). At the population level, density of contacts is correlated with COVID transmission potential (3). Congregate housing environments and related settings with dense human social interactions present ongoing mechanisms for transmission of SARS-CoV-2.

Cruise ship environments are one high-density setting in which COVID transmission dynamics and control measures require further study. Approximately, 29 million people took ocean cruises in 2018; prior to the emergence of COVID-19, 32 million people worldwide were projected to take an ocean cruise in 2020 (4). There have been many outbreaks of infectious diseases on cruise ships in the past — notably respiratory illness outbreaks and norovirus gastroenteritis (5–7). With the emergence of COVID-19, ship outbreaks have become a significant concern. There have already been several COVID-19 outbreaks on cruise ships, aircraft carriers, and cargo ships (8–12).

The outbreak on Diamond Princess cruise ship has been described in substantial detail (12–15). Since it started prior to imposition of widespread social distancing in most locations, the outbreak has been valuable in characterizing SARS-CoV-2 epidemiology (14). On January 20th, 2020 the Diamond Princess departed for a 14-day cruise from Yokohama, Japan (12). On January 25th, a passenger disembarked in Hong Kong and tested positive for COVID 6 days later (16). The Diamond Princess was then quarantined in Yokohama starting on February 5th. Guests and crew were regularly tested and those who tested positive were removed from the ship (12). Starting February 16th, with priority given to people at higher risk for COVID complications, guests who tested negative were allowed to voluntarily disembark and carry out the remainder of their quarantine at a non-medical facility on land (16). On February 23rd, remaining guests were released from the ship in phases to be repatriated, where they were asked to carry out an additional 14-day quarantine (12, 16). Crew served an additional 14-day quarantine after the departure of passengers (12, 16). As of March 27th, 712 of the 3,711 passengers had tested positive for COVID-19, including 311 of whom were asymptomatic at the time of testing and 9 who died (12). This outbreak was key in the early estimate of the fraction of COVID cases who remained asymptomatic, at approximately 18% overall.

Mathematical models have greatly contributed to our understanding of COVID-19 transmission dynamics and optimal strategies for disease control. Many modeling studies have been completed, with most using a traditional ordinary differential equation (compartmental) mathematical framework (17). Because of its assumptions about a large, homogenous, well-mixed population, this framework is limited in representing smaller, dense social settings where the underlying contact networks are highly structured. Rocklöv et. al., for example, was a compartmental model of a simplified stratified ship population (distinguishing ship crew from guests) that estimated empirical transmission parameters on the Diamond Princess (13). However, this model did not represent the full heterogeneity of contact patterns on the ship and did not consider any counterfactual intervention scenarios.

In this study, we developed a dynamic network-based transmission model for COVID-19 on the Diamond Princess. Our two aims were to: 1) characterize the scope and directionality of SARS-CoV-2 transmission aboard the ship given the control events that occurred; and 2) to estimate the epidemiological impact of counterfactual COVID control and prevention strategies. These findings may inform interventions for COVID-19 on future ship sailings, as well as outbreak control in environments with similar high-density contact patterns.

## METHODS

We used a network-based model of infectious disease dynamics to represent the transmission and natural history of SARS-CoV-2 infection across the Diamond Princess cruise ship. This model was built and simulated with the EpiModel software platform (18). This uses the statistical framework temporal exponential random graph models (TERGMs) to estimate and simulate dynamic contact networks based on generative models for network data (19). Our model simulated the individual passengers and crew on the Diamond Princess before and after the major control efforts were implemented to contain the infection spread (Day 15 of the outbreak). The model scenarios were simulated for a period of one month in daily time steps.

### Network Structure

Our model uniquely represented the individuals on the Diamond Princess ship, classified by passenger and crew status (see Figure 1). Following documented records, there were 2666 passengers and 1045 crew, with type represented as a categorical nodal attribute (13, 16). Age was represented as a continuous attribute, with initial distributions drawn from empirical distributions: passengers averaged 69 years old (interquartile range: 62–73) and crew averaged 36 years old (interquartile range: 29–43) (14).

**Figure 1.**
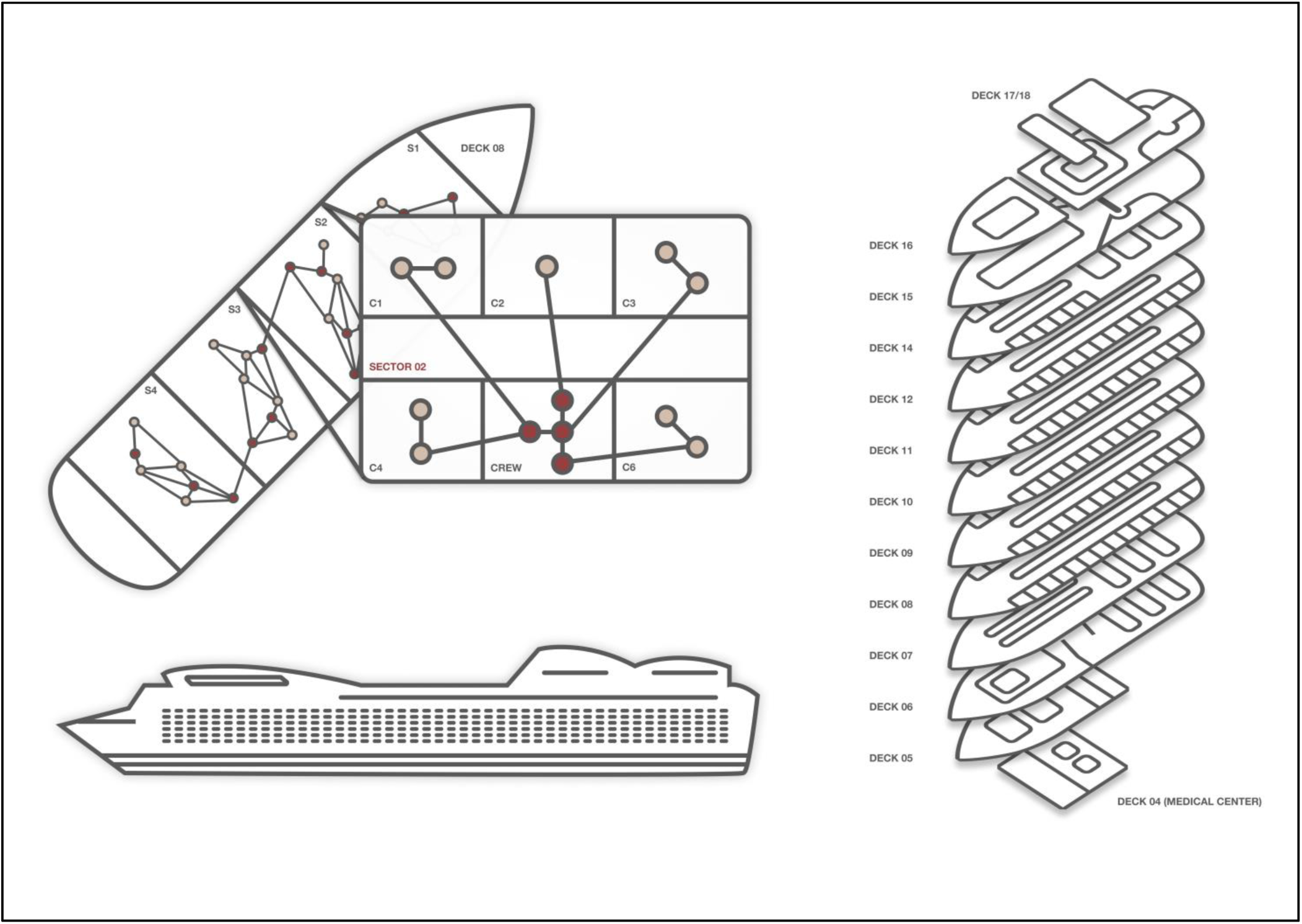
This model schematic represents the structural features of the dynamic contact networks within the ship after the lockdown event was imposed. The larger ship is divided into decks, and on each deck, there are multiple sectors, and within sectors passenger cabins and crew quarters. Networks are comprised of passenger-passenger, passenger-crew, and crew-crew contacts. After lockdown, all passenger-passenger contacts are limited to cabinmates, and crew contacts are substantially reduced to mostly within sector contacts.

Passengers were assigned to one of 1337 cabins on the ship, resulting in an average cabin occupancy of 1.99 passengers. Cabins were then grouped with 10 sectors, which were mostly (but not entirely) self-contained units within the ship that consisted of passenger cabins and assigned crew members. Before control measures were implemented (Day 15 of outbreak), passengers were able to make continued and repeated contacts with their own cabin mates, as well as ongoing contact with random other passengers and crew members both within and outside their cabin sector. The average daily passenger-passenger degree was 5 and passenger-crew degree was 8, based on other models and reports of contact levels and mixing across the ship in a non-outbreak setting (13, 20). Crew members also concentrated the majority of their contacts in their sector prior to network isolation, but also traveled (and thus made contacts) freely across the ship. The average daily crew-crew member degree was 10.

After network lockdown on Day 15, severe restrictions to network degree and cross-sector mixing were simulated, following the empirical characterization of contact isolation (16). This included individuals being confined within their passenger cabins (therefore, making other passenger contacts only with their cabinmates), and crew with limited passenger contact for daily meal and cabin cleaning services. Crew were also constrained to making nearly all their contacts (98%) within their own ship sector to reflect the mobility restrictions imposed upon network lockdown. We assumed within lockdown that the within-dyad contact intensity was 5-fold higher for passenger-passenger contacts compared to passenger-crew and crew-crew contacts to account for the higher frequency of exposure within cabins for passenger-passenger contacts.

We represented these evolving networks using a multi-layer dynamic network approach with TERGMs to simulate the complex interactions that varied by person type and time. A total of 6 TERGMs were fit to the network degree distribution statistics above: one for each contact type interaction (passenger-passenger, passenger-crew, and crew-crew), doubled for before and after the network lockdown on Day 15 that fundamentally altered the network structure. The TERGMs were estimated and simulated using standard MCMC-based fitting procedures (19, 21), and then diagnosed by comparing the simulated network data against the input data points.

### COVID Transmission and Progression

Our model represented pathogen transmission and disease progression following common COVID-19 modeling approaches (17). This SEIR framework allowed for infected persons to stochastically transition from susceptible to exposed (latent) stages upon infection (Supplemental Figure 1). Persons then transitioned to either a symptomatic or asymptomatic pathway for the infectious period before recovering. Following estimates from the Diamond Princess, 25–76% of persons entered into the symptomatic infectious pathway, with the probability of symptoms positively correlated with age (17). Asymptomatic infected persons had 50% the transmission potential compared to those with symptomatic infection (22).

For persons in the symptomatic pathway, states were divided into a pre-clinical infectious followed by a clinical infectious stage. In the base model, all persons in the clinical infectious stage reduced their contact intensity by 90% after the outbreak was recognized (Day 15). We implemented a mortality process, with general age-specific mortality rates following standardized age-specific mortality data (23), with a multiplier for excess COVID-related mortality within the clinical infectious state (representing the most severe disease) only. This multiplier was calibrated to reproduce the number of COVID-19 deaths at one month.

Widespread screening was imposed, with screening rates stratified by symptomatic status, following empirical diagnostic patterns on the Diamond Princess (16). In the base scenario, widespread screening was initiated on Day 15, with higher screening rates for symptomatic cases. We calibrated the stratified daily rates to reproduce the daily case count on the ship. The calibrated screening rates resulted in approximately 3,100 cases screened over the month, with 634 positive cases at one month (see Figure 2). We used a PCR sensitivity of 80% (24), and assumed that diagnosed persons reduced their contact intensity by 90% (similar to the reduction for symptomatic cases).

**Figure 2.**
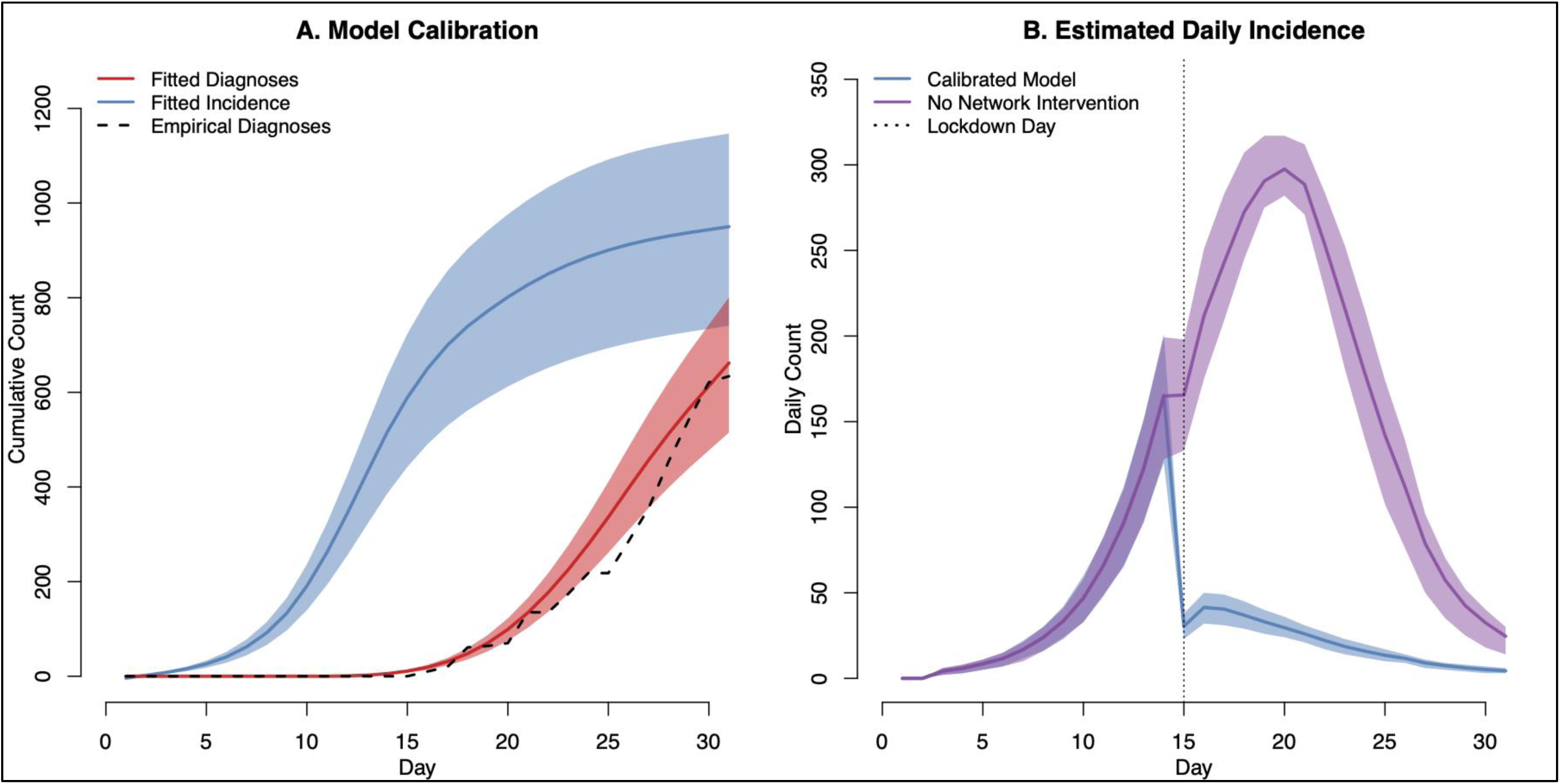
Panel A (left) shows the results of the primary model calibration, which was to the cumulative diagnoses of COVID-19 that occurred through month 1 on the ship. The primary diagnosis efforts were initiated on Day 15. Model calibration parameters included the symptomatic and asymptomatic diagnosis rates per day and the transmission probability per exposure. The total incidence of disease consistent with this calibration is shown in blue. Panel B (right) shows the daily incidence of all (diagnosed and undiagnosed) new COVID-19 cases in the calibrated model, and then in a model scenario in which there was no network lockdown intervention. The network lockdown date on Day 15 is shown in a vertical line. Both panels display the median (dark line) and interquartile range (light bands) from 1000 simulations of the calibrated model.

### Intervention Scenarios

We modeled three categories of prevention and control interventions: behavioral, clinical, and biological. Control interventions were those imposed in response to the outbreak on the ship (i.e., on or after Day 15) whereas prevention interventions were counterfactual scenarios that could reduce SARS-CoV-2 transmission before an outbreak (i.e., what might be useful for future cruises).

For behavioral interventions, we investigated the impact of the timing of the network lockdown and the intensity of the contact reduction of each of the sub-networks (passenger-passenger, passenger-crew, and crew-crew). Outcomes were the cumulative incidence and mortality at one month. The network lockdown time was varied by implementing the restricted network at counterfactual days across the month, relative to the Day 15 base scenario. Our question was whether imposing the lockdown at an earlier day could have prevented a major outbreak on the ship. We also investigated whether reducing the intensity of contact (conditional on a network dyad existing) could lower overall cumulative incidence, or incidence of cases with a particular contact directionality. This contact intensity reduction was implemented by reducing the per-dyad daily contact rate by a relative amount, where the calibrated base scenario was 100%. This contact intensity reduction was implemented in either in tandem with the Day 15 network lockdown or on Day 1 in the absence of a lockdown.

Clinical interventions consisted of varying the timing of mass screening on the ship, and the intensity of case isolation after a positive diagnosis. We first evaluated, from a control perspective, how varying the intensity of case isolation would reduce the cumulative incidence (the base scenario assumed a 90% reduction). This was evaluated at Day 15, with and without network lockdown, with the contact intensity reduced starting on Day 15. We next estimated, from a prevention perspective, how varying the timing of mass screening from Day 1 to never, would impact the cumulative incidence at one month. Bivariate sensitivity analyses further explored the relationship between screening start date, diagnosis-based contact intensity reduction, and PCR test sensitivity on cumulative incidence.

Biological interventions were simulated by varying the intensity of personal protective equipment (PPE; e.g., face masks) use across the behavioral and clinical interventions. This was implemented in different counterfactual scenarios in which PPE was uniformly used or not used by everyone on the ship in order to understand the total causal effect of PPE use as a ship-wide intervention. We assumed that PPE was used for passenger-crew and crew-crew contact types, but not passenger-passenger contacts after lockdown (because of the difficulty in maintaining these interventions within passenger cabins). This involved modifying the force of infection per contact by a reduction of 40% in the PPE scenarios (25).

### Calibration, Simulation, and Analysis

Our model was calibrated to the cumulative diagnosis curve through 1 month as observed and reported on the Diamond Princess (16). Free parameters for model calibration were: the daily screening rate (stratified by symptomatic status), which was allowed to vary in 5-day blocks to avoid overfitting; the intensity for passenger-passenger contacts per day; and the probability of infection per contact. The base calibrated model scenario implemented network lockdown, clinical interventions, and PPE use starting on Day 15. We fit the model to cumulative diagnoses per day, the total number of screening tests at one month, and the total number of deaths at one month. Figure 2 shows the results of this calibration relative to data.

For each reference and counterfactual scenario, we simulated the model 1000 times and summarized the results with medians and 95% simulation intervals. Outcome measures were the cumulative incidence of COVID-19 and the mortality at 1 month. We also quantified the directionality of transmissions by tracking who infected whom by person type (passenger versus crew) on the ship. Outcomes of the number and percent of infections (or deaths) averted were used to compare the cumulative incidence (or mortality) in an experimental scenario relative to the base scenario.

## RESULTS

Figure 2 shows the results of the model calibration and timing of the network-related control measures on the Diamond Princess ship. Day 1 represents January 20, and Day 15 represents March 4. The empirical number of positive diagnosed cases at one month was 634, compared to the fitted model of 647 (interquartile range [IQR]: 504, 785). The total incidence consistent with this calibration, which includes undiagnosed cases and false-negative cases (due to imperfect PCR sensitivity), was 948 (IQR: 739, 1146). Panel B shows that the daily total incidence in the calibrated model that includes empirical network lockdown, compared to a model in which no network intervention occurred. In the calibrated model, peak incidence occurred on Day 14, with 165 cases (IQR: 126, 201). In the no-intervention model, peak incidence occurred on Day 20, with daily 298 cases (IQR: 282, 317). In the calibrated model, 66.1% of total cumulative cases had occurred by Day 15 whereas in the no-intervention model, only 21.7% had occurred by Day 15.

Figure 3 shows the results of the primary network intervention, stratified by PPE use starting at the network lockdown. Corresponding numerical results are provided in Supplemental Table 1. The density of cumulative incidence across the simulations in each scenario is visualized given the model stochasticity. The variability of outcomes within scenario is partially driven by the random seeding of infection and the case clustering within passenger cabins.

**Figure 3.**
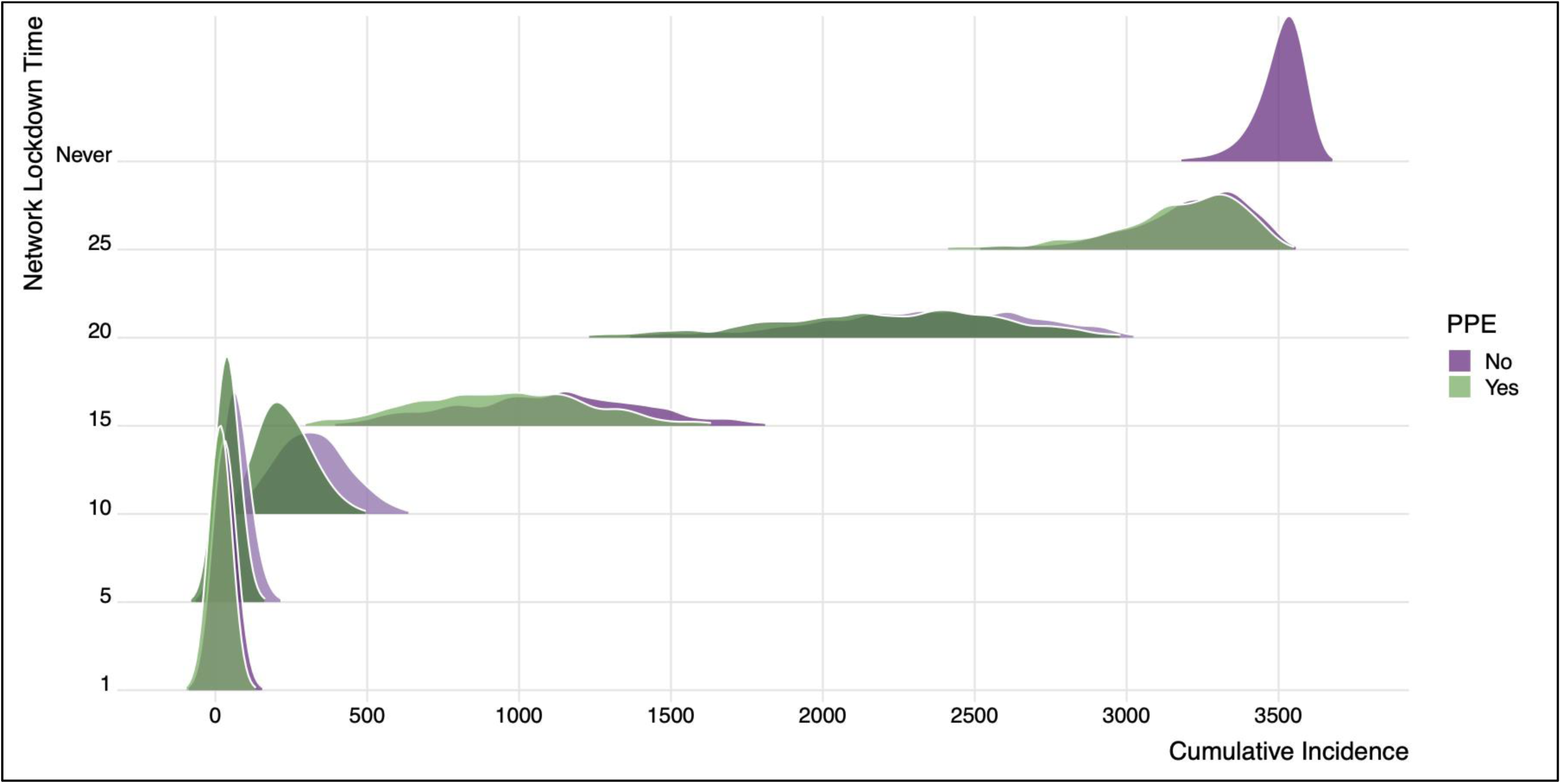
This figure shows the distribution of cumulative incidence in the model scenarios that vary the network lockdown time, with and without personal protective equipment (PPE). The base (calibrated) model corresponds to Day 15, when the actual control efforts were implemented. If the network lockdown had never been implemented, over 3500 cases would have been projected. If the lockdown had occurred one Day 1, fewer than 20 cases would be projected. This figure shows the empirical distribution of cumulative incidence across 1000 simulations of each scenario.

In the base scenario, there were an estimated 948 infections and 10 COVID-related deaths. Implementing the network lockdown with PPE 10 days earlier (Day 5) would result in 909 fewer infections and no COVID-19 deaths. Implementing the network lockdown 10 days later (Day 25) was projected to result in 2224 more cases and 20 more COVID deaths. The overall impact of PPE was minimal in these scenarios, conditional on the network lockdown time. The cumulative incidence if the network lockdown had occurred on the same day as the base model (Day 15) but no PPE were ever used would be 1113 cases (152 more than the base scenario with PPE) and 11 deaths (1 more than the base scenario). The impact of PPE was minimal here because of the high network degree and contact intensity within each network link, overwhelming the per-contact reduction in the force of infection from PPE, and the directionality of transmission.

Supplemental Table 2 shows the directionality of transmission events in the base calibrated model with network lockdown and PPE at Day 15, and also in a counterfactual scenario in which there was no network lockdown but PPE use was initiated on Day 1. In the base model, 59% of cumulative cases (551 out of 934) were passenger to passenger transmissions, compared to 17% that were passenger to crew, 13% that were crew to passenger, and 10% that were crew to crew. Control-based interventions aimed at further reducing the contact intensity within each contact type are shown as counterfactuals. If implemented after network lockdown, contact intensity reductions could avert up to an additional 190 cases (in the scenario in which passenger-crew contact intensity were reduced by 100%). This directional contact intensity reduction prevents cases directly (from passengers to crew and crew to passengers) but also indirectly in the other two sub-networks (passenger to passenger and crew to crew). With contact intensity reductions implemented at Day 1 (as a prevention intervention in the absence of network lockdown), even extreme levels of passenger-passenger contact intensity reduction (which would not be realistic on a ship) failed to avert the same number of cases as the network lockdown scenario implemented on Day 1. These scenarios highlight that passenger-passenger contact continues to be the dominant mode of transmission.

Supplemental Table 3 shows the results of the intervention of using diagnoses that prompt case isolation, using the control-based perspective in which the interventions were implemented on Day 15 after screening ramped up. Base models assumed that diagnosis resulted in a 90% reduction in contact intensity across all three contact networks. Even if case isolation were completely relaxed, with no case isolation starting with any diagnoses on Day 15, there would be a minimal impact on cumulative incidence and mortality. The same effects are observed in the set of scenarios in which no network lockdown occurred, but in which case isolation still beginning on Day 15.

Mechanistic reasons for the minimal impact of case isolation are explored in the next scenario set (Figure 4 and Supplemental Table 4) that implemented case isolation from a prevention perspective. These scenarios varied the timing of mass screening of all persons. The Day 1 timing scenario is consistent with pre-screening of all passengers and crew on the ship, as we (optimistically) assumed that results delivery and case isolation occurred on the same day. These scenarios further assumed that no network lockdown or other general contact restriction occurred, in order to represent a natural ship environment without contact restrictions. These scenarios also assumed that diagnosis-based and symptoms-based case isolation was complete (100% of cases diagnosed or with COVID symptoms are isolated immediately).

**Figure 4.**
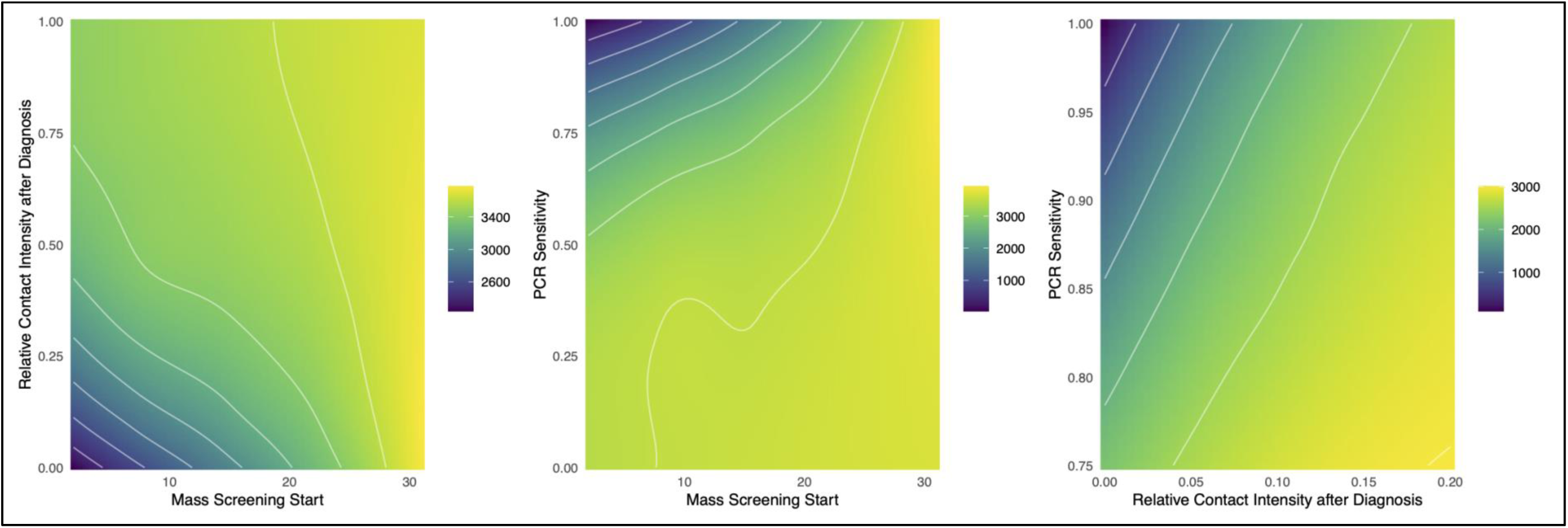
This figure shows the projected total cumulative incidence at 1 month across scenarios for COVID-19 control based on case isolation after PCR diagnosis. Panel A (left) shows the impact on cumulative incidence based on timing of mass (asymptomatic) screening against intensity of isolation of diagnosed cases (where 0 = full isolation and 1 = no isolation); this highlights that incidence is minimized by screening begins immediately and is associated with complete case isolation (mean incidence in bottom left = 2244). Panel B (middle) shows the impact on cumulative incidence based on timing of mass screening against PCR sensitivity, where the base sensitivity is 80% and the diagnosed isolation intensity is 0; this highlights that incidence is minimized when screening begins immediately and PCR sensitivity is perfect (mean incidence in top left = 0). Panel C (right) shows the impact on cumulative incidence based on diagnosed isolation intensity and PCR sensitivity, with mass screening starting at Day 1; this highlights that incidence is minimized when isolation intensity is highest and PCR sensitivity is perfect (mean incidence in top left = 0).

If mass screening and case isolation were to occur on Day 1, the projected incidence would be 2286 cases, which was 1404 fewer than the scenario in which this prevention strategy were never implemented. Projected cumulative mortality in this scenario was 7 cases (95% SI: 0, 24). PPE use contributed more significantly to prevention in these scenarios than in the network lockdown scenarios in Figure 1 because it would be used for the entire month and a larger fraction of contacts were outside the passenger cabins (PPE use was not represented within cabins). However, even with complete PPE use and immediate diagnosis-based case isolation, the cumulative incidence was 1630 cases and 5 deaths.

Figure 4 highlights the relationships between the prevention mechanisms for this diagnosis-based case isolation. Panels A (left) and B (middle) demonstrate that the optimal timing of mass screening was as soon as possible. Cumulative incidence was further minimized when relative contact intensity after diagnosis trended towards zero (complete isolation). However, in this case the minimal cumulative incidence (in the bottom left scenario) was still 2244. Panel B shows that cumulative incidence was further minimized in an early mass screening scenario with increased PCR sensitivity. In comparison with a base model PCR sensitivity of 80% (resulting in 20% of true-positive cases remaining undiagnosed), a test sensitivity approaching 100% would minimize incidence, assuming complete contact isolation after diagnosis. Panel 3 shows that both case isolation must be complete (relative contact intensity = 0) and PCR sensitivity must be 100% to achieve complete outbreak prevention (in absence of a network lockdown). The expected cumulative incidence in these ideal scenarios was zero.

## DISCUSSION

In our model of SARS-CoV-2 on a cruise ship, we found that only extreme restrictions in social contact or idealistic clinical response strategies could fully avert a major COVID outbreak. Contact network changes associated with adequate outbreak prevention were the restriction of passengers to their cabins, with limited passenger-crew contacts. Clinical response strategies required for outbreak prevention included early mass screening with an ideal PCR test (100% sensitivity) and immediate case isolation upon diagnosis. Without these behavioral and clinical interventions, it was projected that hundreds of COVID-19 cases (with tens of associated deaths) would occur on a ship the size of the Diamond Princess. Ultimately, the public health costs of COVID-19 on a cruise ship environment are likely greater than the leisure benefits. Public health restrictions on activities like these should be considered until longer-term effective solutions such as a SARS-CoV-2 vaccine become widely available.

The driving reason for the extreme interventions necessary for outbreak prevention is the overwhelming force of infection on a high-density setting like a cruise ship. Outbreaks in congregate living and other high-density contact environments have been observed, including in prisons, nursing homes, and university dormitories (26, 27). Outbreak control has been challenging in these settings because of the need to maintain ongoing social distancing in environments where that is often infeasible. Mathematical models have also identified the substantial clinical resources needed for further disease control there (28). The primary effective response in these settings therefore is to implement something like our network lockdown scenario, by restricting contacts to the minimum, and reducing the overall intensity of contacts within these settings (e.g., releasing prisoners or reducing campus population density). Outbreaks on cruise ships will likely continue without substantial contact restriction that may be at odds with the intended purposes of these vessels.

A methodological strength of our model is its dynamic network framework. This allowed for microsimulation of persons over time as their contact networks and disease dynamics co-evolve. This framework avoids the mass action assumptions of ordinary differential equation models violated by these small-scale but high-density ship environments (29). The stochastic nature of network models also allowed for projection of the range of outcomes within scenarios, especially useful for estimating the probability of outbreaks. Several network-based models for COVID-19 have been developed (7, 20, 30, 31), including models for social contact restriction in community-based settings and ship environments. One strength of our specific network modeling approach with TERGMs is the representation of dynamic networks in which both the node set and edge set are responsive to epidemic dynamics in statistically principled ways (32). These network models are also parametrized with representations of contact patterns on the ship rather than idealized (e.g., small-world) networks.

### Limitations

The primary limitation of our study was the assumption about network contact patterns on the ship prior to the network lockdown. While the contact patterns on the Diamond Princess were reasonably well-characterized after the contact restrictions, behavior on the ship prior to that point was been less estimated. Additionally, what is considered an effective contact for respiratory diseases requires behavioral and biological assumptions to decompose the elements of R_0_. While our models require more assumptions than differential equation models, our model outputs were also consistent with the broad population outcomes from those models. For example, we were able to represent the underlying transmission potential projections of the earlier Diamond Princess models and analyses (13, 14). Further network data on the unique contact patterns within high-density settings like ships would be greatly informative to future network modeling research.

## Conclusions

Resuming cruise ship activities before a long-term COVID prevention solution becomes available may be inadvisable without fundamentally changing the nature of activities on these vessels. Our findings have implications for other high-density social contact settings in which the multi-layer network contact patterns may drive the high force of infection at multiple hierarchies of contact. In settings where severe contact restriction (e.g., network lockdown) is infeasible, substantial clinical resources (mass screening with rapid but high-sensitivity diagnostic results delivery and case isolation) would be needed for complete outbreak prevention.

## Data Availability

No primary data were used in this study. Model code is available at https://github.com/EpiModel/COVID-CruiseShip.

