## Supplemental Appendix for "Dynamic Network Strategies for SARS-CoV-2 Control on a Cruise Ship"

### 1 INTRODUCTION

This supplementary technical appendix describes the mathematical model structure, parameterization, and statistical analysis of the accompanying paper in further detail.

#### 1.1 Model Framework

The mathematical models for SARS-CoV-2 transmission dynamics presented in this study are network-based transmission models in which uniquely identifiable relational contact dyads were simulated and tracked over time. This contact structure is represented through the use of temporal exponential-family random graph models (TERGMs). On top of this dynamic network simulation, the epidemic model represents demography (exits, and aging), interhost epidemiology (disease transmission), intrahost epidemiology (disease progression), and clinical epidemiology (disease diagnosis and treatment and prevention interventions). Individual attributes related to these processes are stored and updated in discrete time over the course of each epidemic simulation.

#### 1.2 Model Software

The models in this study were programmed in the R and C++ software languages using the *EpiModel* [http://epimodel.org/] software platform for epidemic modeling. *EpiModel* was developed by the authors for simulating complex network-based mathematical models of infectious diseases.(1) *EpiModel* depends on *Statnet* [http://statnet.org/], a suite of software in R for the representation, visualization, and statistical analysis of complex network data.(2)

*EpiModel* allows for a modular expansion of its built-in modeling tools to address novel research questions. We have developed a set of extension modules into a software package called *EpiModelCOVID*. This software is available for download, along with the scripts used in the execution of these models. The tools and scripts to run these models are contained in two GitHub repositories:

- [http://github.com/EpiModel/EpiModelCOVID] contains the general extension software package. Installing this using the instructions listed at the repository homepage will also load in *EpiModel* and the other dependencies.
- [http://github.com/EpiModel/COVIDCruiseShip] contains the scripts to execute the models and to run the statistical analyses provided in the manuscript.

#### 1.3 Core Model Specifications

We started with a network size of 3711 persons on the Diamond Princess Cruise Ship. Age was represented as a continuous attribute, with initial distributions drawn from empirical distributions: passengers averaged 69 years old (interquartile range: 62–73) and crew averaged 36 years old (interquartile range: 29–43). The network size was allowed to decrease with departures related to mortality. We used a two-stage simulation framework, first calibrating the model to diagnosed cases on the ship (Stage 1), and then simulating the reference and counterfactual intervention scenarios for 30 days in most scenarios. The time unit used throughout the simulations was one day. Unless otherwise noted, all rate-based parameters listed below are to be interpreted as the rate per day and all duration-based estimates are to be interpreted as the duration in days.

### 2 NETWORKS OF SOCIAL CONTACTS

We modeled networks of three interacting types of network contacts relations: passenger to passenger, passenger to crew, and crew to crew. We first describe the methods conceptually, including the parameters used to guide the model and their derivation, and then present the formal statistical modeling methods. Consistent with our parameter derivations, all contacts are defined as those in which respiratory exposure is expected to occur at least once.

#### 2.1 Conceptual Representation of Networks

Our modeling methods aim to preserve certain features of the cross-sectional and dynamic network structure as observed in our primary data, while also allowing for mean contact durations to be targeted to those reported for different groups and relational types. Our methods do so within the context of changing population size (due to deaths and departures from the population) and changing composition by attributes such as age.

This model involved representing three types of contact networks at two distinct time points. The three networks represented passenger-passenger, passenger-crew, and crew-crew contacts. We modeled these as three overlapping networks with a shared node set but differing edges (instead of one large network) to provide maximal flexibility in model parameterization and intervention design. The three modeled networks were also doubled to represent a “pre-lockdown” and “post-lockdown” composition. Although the core network features remained the same, the density and determinants of the network structure varied between these time points.

Conceptually, the 6 network models are defined as follows:

- Passenger-Passenger Network
    - Pre-Lockdown
        - Contacts between passengers, with most contacts occurring with passenger in same cabin. Average daily degree of 5.
        - Contacts prohibited with crew members in this network.
    - Post-Lockdown
        - All passenger-passenger contacts limited to within-cabin. Same within-cabin daily degree (average degree of 1) but no contacts with passengers outside of cabin.
        - Contacts prohibited with crew members in this network.
- Passenger-Crew Network
    - Pre-Lockdown
        - Contacts between passengers and crew, with an average daily degree of 8.

        - 50% of contacts restricted to passenger-crew pairs within same ship sector.
        - No contacts within person type permitted.
    - Post-Lockdown
        - Contacts limited to 2 daily visits to each cabin (for cleaning and meal services)
        - 98% of contacts restricted to passenger-crew pairs within the same ship sector.
- Crew-Crew Network
    - Pre-Lockdown
        - Contacts between crew, with an average daily degree of 10.
        - Contacts prohibited with passengers in this network.
    - Post-Lockdown
        - Contacts between crew, with an average daily degree of 2.
        - 98% of contacts restricted to crew-crew pairs within same ship sector.
        - Contacts prohibited with passengers in this network.

Algorithmically, we implemented the swap of network models from pre-lockdown to post-lockdown by implementing a deterministic time step at which the appropriate network model was selected as input for the simulation.

#### 2.2 Statistical Representation of Contact Networks

Exponential-family random graph models (ERGMs) provide a foundation for statistically principled simulation of local and global network structure given a set of target statistics from empirical data. Social contacts were modeled using modeled using cross-sectional ERGMs.(3) This allowed for more flexibility in relational dissolution than temporal ERGMs, in which this is a stochastic process. Repeated contacts within the same dyads (e.g., passenger cabinmates) were allowed through mixing constraints in the cross-sectional network composition.

Formally, our statistical models for relational dynamics can be represented as a set of six equations for the conditional log odds (logits) of relational existence at time *t*:

$$logit\left(P\left(Y_{ij,t<15}=1\middle|\,Y_{ij,t}^{C}\right)\right)=\theta_{pp}{}'\partial\left(g_{pp}(y)\right)$$ passenger-passenger, pre-lockdown

$$logit\left(P\left(Y_{ij,t\geq 15}=1\middle|\,Y_{ij,t}^{C}\right)\right)=\theta_{pp}{}'\partial\left(g_{pp}(y)\right)$$ passenger-passenger, post-lockdown

$$logit\left(P\left(Y_{ij,t<15}=1\middle|\,Y_{ij,t}^{C}\right)\right)=\theta_{pc}{}'\partial\left(g_{pc}(y)\right)$$ passenger-crew, pre-lockdown

$$logit\left(P\left(Y_{ij,t\geq 15}=1\middle|\,Y_{ij,t}^{C}\right)\right)=\theta_{pc}{}'\partial\left(g_{pc}(y)\right)$$ passenger-crew, post-lockdown

$$logit\left(P\left(Y_{ij,t<15}=1\middle|\,Y_{ij,t}^{C}\right)\right)=\theta_{cc}{}'\partial\left(g_{cc}(y)\right)$$ crew-crew, pre-lockdown

$$logit\left(P\left(Y_{ij,t\geq 15}=1\middle|\,Y_{ij,t}^{C}\right)\right)=\theta_{cc}{}'\partial\left(g_{cc}(y)\right)$$ crew-crew, post-lockdown

where:

- $Y_{ij,t}$ = the relational status of persons *i* and *j* at time *t* (1 = in relationship/contact, 0 = not).

- $Y_{ij,t}^{C}$ = the network complement of *i,j* at time *t*, i.e. all relations in the network other than *i,j.*
- $g(y)$ = vector of network statistics in each model (the empirical statistics defined in the tables above).
- $\theta$ = vector of parameters in the model.

For $g(y)$ and $\theta$, the subscript indicates the contact network type. For $Y_{ij,t}$ the subscript differentiates the pre- and post-lockdown times corresponding to day 15. The recursive dependence among the relationships renders the model impossible to evaluate using standard techniques; we use MCMC in order to obtain the maximum likelihood estimates for the $\theta$ vectors given the $g(y)$ vectors.

Converting the statistics into our fully specified network models consists of the following steps:

1. Construct a cross-sectional network of 3711 persons with no contacts (an empty network).
2. Assign persons demographics (age) based on ship census data, as well as a passenger type (corresponding to passenger or crew). For passengers, individuals were assigned a cabin number (and thus cabin mate). Both passengers and crew were assigned a sector on the ship to which the cabin and crew were assigned.
3. Calculate the target statistics (i.e., the expected count of each statistic at any given moment in time) associated with the terms in the existence model.
4. Estimate the coefficients for the existence model that represent the maximum likelihood estimates for the expected cross-sectional network structure.

Steps 1–4 occur within the *EpiModel* software, and use the ERGM methods therein. They are completed efficiently by the use of an approximation in Step 4 (4). During the subsequent model simulation, we use the method of Krivitsky (5) to adjust the coefficient for the first term in each model at each time step, in order to preserve the same expected mean degree (relationships per person) over time in the face of changing network size and nodal composition. At all stages of the project, simulated partnership networks were checked to ensure that they indeed retained the expected cross-sectional structure and relational durations throughout the simulations.

#### SUPPLEMENTAL FIGURES

**Supplemental Figure 1.** COVID-19 transmission and disease progression is represented as transitions from an exposed latent state to either a symptomatic (clinical) or asymptomatic (subclinical) pathway. Transmissibility is reduced within the asymptomatic pathway. Disease-induced mortality occurs within the infectious, symptomatic state only.

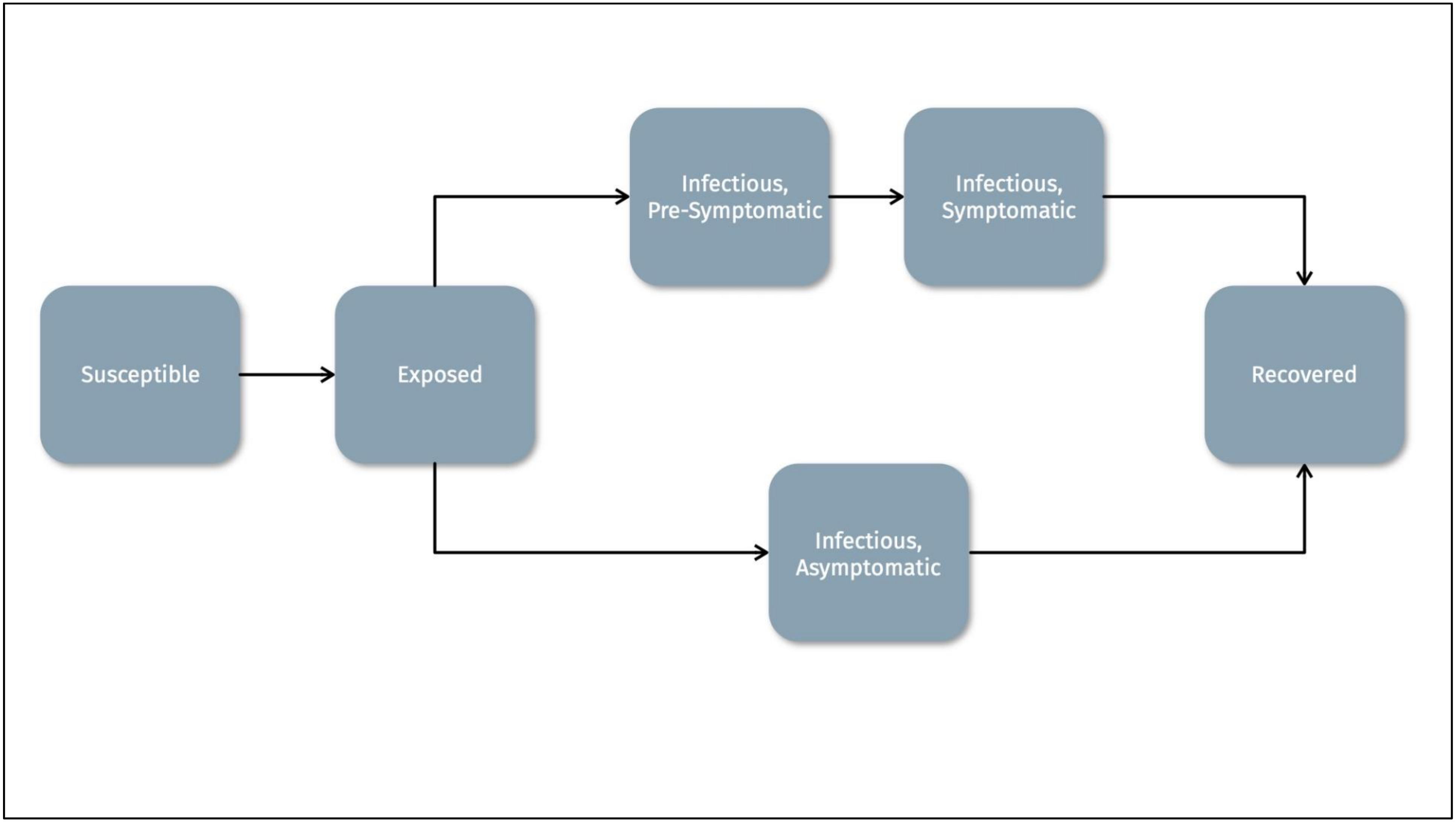

#### SUPPLEMENTAL TABLES

**Supplemental Table 1.** Primary Model Parameters

| Parameter | Value | Source |
|---|---|---|
| Transmission probability per contact | 11% | Kraay 2020 (6); Fitted |
| Relative reduction in transmission with PPE | 40% | Chu 2020 (7) |
| Passenger-Passenger Daily Mean Degree, Pre-Lockdown | 5 | Rocklov 2020 (8); Xue 2020 (9) |
| Passenger-Passenger Proportion of Network Degree within Cabin, Pre-Lockdown | 20% | Moriarty 2020 (10); Assumed |
| Passenger-Passenger Daily Mean Degree, Post-Lockdown | 1 | Moriarty 2020 (10); Rocklov 2020 (8) |
| Passenger-Passenger Proportion of Network Degree within Cabin, Post-Lockdown | 100% | Moriarty 2020 (10); Assumed |
| Passenger-Crew Daily Mean Degree, Pre-Lockdown | 8 | Rocklov 2020 (8); Xue 2020 (9) |
| Passenger-Crew Proportion of Network Degree within Same Sector, Pre-Lockdown | 50% | Moriarty 2020 (10); Assumed |
| Passenger-Crew Daily Mean Degree, Post-Lockdown | 2 | Moriarty 2020 (10); Rocklov 2020 (8) |
| Passenger-Crew Proportion of Network Degree within Same Sector, Post-Lockdown | 98% | Moriarty 2020 (10); Assumed |
| Crew-Crew Daily Mean Degree, Pre-Lockdown | 10 | Rocklov 2020; Xue 2020 |
| Crew-Crew Daily Mean Degree, Post-Lockdown | 2 | Moriarty 2020; Rocklov 2020 |
| Crew-Crew Proportion of Network Degree within Same Sector, Post-Lockdown | 98% | Moriarty 2020 (10); Assumed |
| Passenger-Passenger within Dyad Exposures | 5 per day | Fitted |
| Passenger-Crew within Dyad Exposures | 1 per day | Fitted |
| Crew-Crew within Dyad Exposures | 1 per day | Fitted |
| Within Dyad Exposure Rate Reduction Following Positive Diagnosis | 0.1 multiplier | Moriarty 2020 (10); Chu (7); Assumed |
| Proportion Symptomatic | 10–19 years: 40%<br>20–29 years: 25%<br>30–39 years: 37%<br>40–49 years: 42%<br>50–59 years: 51%<br>60–69 years: 59%<br>70–79 years: 72%<br>80+ years: 76% | Davies 2020 (11) |
| Duration of Latent Period | 4 days | Davies 2020 (11) |
| Duration of Preclinical Infectious Period | 1.5 days | Davies 2020 (11) |

| | | |
|---|---|---|
| Duration of Clinical Infectious Period | 3.5 days | Davies 2020 (11) |
| Duration of Subclinical Infectious Period | 5 days | Davies 2020 (11) |
| Natural Mortality Rate | Yearly age-specific rate 0 to 100 years | Global Burden of Disease (12) |
| COVID-Related Mortality (Multiplier on natural mortality) | 180 | Fitted |
| PCR Test Sensitivity | 80% | Lopman 2020 (13) |
| Number of Ship Passengers | 2,666 | Moriarty 2020 (10) |
| Number of Crew | 1,045 | Moriarty 2020 (10) |

**Supplemental Table 2.** Impact of Network Isolation Timing, With and Without Personal Protective Equipment (PPE), on COVID-19 Incidence and Mortality at 1 Month

| Scenario | Cumulative Incidence | | | Cumulative Mortality | | |
|---|---|---|---|---|---|---|
| | Total | NIA[1] | PIA[2] | Total | NDA[3] | PDA[4] |
| | *Median (95% SI)* | *Median (95% SI)* | *Median (95% SI)* | *Median (95% SI)* | *Median (95% SI)* | *Median (95% SI)* |
| *Base Isolation Scenario* | | | | | | |
| Day 15 (with PPE) | 948.0 (388.0, 1534.2) | – | – | 10.0 (3.0, 20.0) | – | – |
| *Varying Network Isolation Time (with PPE)* | | | | | | |
| Day 1 | 15.0 (5.0, 32.0) | 931.5 (928.0, 935.0) | 98.4 (98.3, 98.4) | 0.0 (0.0, 1.0) | 10.0 (10.0, 10.0) | 100.0 (100.0, 100.0) |
| Day 5 | 38.0 (12.0, 80.0) | 909.0 (903.5, 914.0) | 96.0 (95.9, 96.1) | 0.0 (0.0, 2.0) | 10.0 (10.0, 10.0) | 100.0 (100.0, 100.0) |
| Day 10 | 221.0 (91.0, 418.1) | 715.0 (704.0, 726.0) | 76.0 (75.4, 76.5) | 2.0 (0.0, 7.0) | 8.0 (8.0, 8.0) | 77.8 (77.8, 78.6) |
| Day 20 | 2225.5 (1328.0, 2866.1) | -1265.2 (-1287.5, -1243.0) | -132.2 (-135.6, -128.6) | 23.0 (11.0, 36.0) | -13.0 (-13.0, -13.0) | -127.3 (-133.3, -122.2) |
| Day 25 | 3205.5 (2410.6, 3465.0) | -2224.0 (-2239.0, -2208.0) | -232.6 (-235.9, -228.8) | 31.0 (18.0, 43.0) | -20.0 (-21.0, -20.0) | -200.0 (-209.1, -196.4) |
| None | 3522.0 (3298.0, 3600.0) | -2557.0 (-2567.5, -2547.0) | -269.4 (-271.6, -267.3) | 32.0 (19.0, 45.0) | -22.0 (-22.0, -21.0) | -212.5 (-220.0, -208.3) |
| *Varying Network Isolation Time (no PPE)* | | | | | | |
| Day 1 | 28.0 (8.0, 67.0) | 918.0 (912.5, 923.0) | 97.0 (96.9, 97.1) | 0.0 (0.0, 2.0) | 10.0 (10.0, 10.0) | 100.0 (100.0, 100.0) |
| Day 5 | 62.0 (19.0, 136.1) | 881.0 (874.5, 887.5) | 93.3 (93.2, 93.5) | 0.0 (0.0, 3.0) | 9.0 (9.0, 10.0) | 100.0 (100.0, 100.0) |
| Day 10 | 315.5 (121.0, 560.1) | 625.5 (614.0, 638.0) | 66.5 (65.8, 67.2) | 3.0 (0.0, 8.0) | 7.0 (7.0, 7.0) | 72.7 (71.4, 75.0) |
| Day 15 | 1112.5 (490.0, 1755.1) | -151.5 (-171.5, -132.5) | -16.0 (-18.4, -13.8) | 11.0 (3.0, 22.0) | -1.0 (-1.0, 0.0) | -6.7 (-9.1, 0.0) |
| Day 20 | 2336.0 (1402.0, 2928.1) | -1380.5 (-1400.5, -1358.5) | -144.0 (-147.4, -140.5) | 23.0 (11.0, 37.0) | -13.0 (-13.0, -13.0) | -129.7 (-133.4, -124.0) |
| Day 25 | 3228.0 (2520.6, 3481.0) | -2254.5 (-2271.0, -2240.0) | -236.0 (-239.4, -232.7) | 31.0 (17.0, 45.0) | -21.0 (-22.0, -21.0) | -209.5 (-216.7, -200.0) |
| None | 3520.0 (3255.9, 3603.0) | -2556.5 (-2566.0, -2547.5) | -269.5 (-271.8, -267.0) | 32.0 (19.0, 45.0) | -22.0 (-22.0, -21.0) | -214.8 (-222.2, -210.0) |

[1] Number of infections averted relative to base scenario

[2] Percent of infections averted relative to base scenario

[3] Number of COVID-related deaths averted relative to base scenario

[4] Percent of COVID-related deaths averted relative to base scenario

**Supplemental Table 3.** Directionality of Transmission and Contact Intensity Reductions, with Day 15 Network Lockdown and PPE, on COVID-19 Incidence at 1 Month

| Scenario | **Total** | **Passenger to Passenger** | **Passenger to Crew** | **Crew to Passenger** | **Crew to Crew** |
|---|---|---|---|---|---|
| | Cumulative Incidence | Cuml. Incid. | Cuml. Incid. | Cuml. Incid. | Cuml. Incid. |
| | *Median (95% SI)* | *Median (95% SI)* | *Median (95% SI)* | *Median (95% SI)* | *Median (95% SI)* |
| **With Contact Intensity Reductions, Network Lockdown, and PPE at Day 15** | | | | | |
| *Base Scenario* | | | | | |
| No Intensity Reduction | 933.5 (366.0, 1556.2) | 551.0 (213.9, 941.0) | 163.0 (66.0, 265.0) | 124.0 (46.0, 211.0) | 93.0 (33.0, 175.0) |
| *Varying Passenger-Passenger Contact Intensity* | | | | | |
| 50% Reduction | 862.5 (353.9, 1454.0) | 488.0 (203.9, 843.0) | 155.0 (67.0, 257.0) | 124.5 (47.0, 216.0) | 93.5 (29.0, 174.0) |
| 90% Reduction | 765.5 (316.9, 1348.0) | 401.0 (164.9, 727.0) | 145.5 (63.0, 248.0) | 122.0 (44.0, 214.0) | 90.0 (31.0, 173.0) |
| 100% Reduction | 749.0 (297.9, 1255.1) | 381.0 (155.9, 677.0) | 147.5 (61.0, 241.0) | 126.0 (44.0, 208.0) | 93.0 (32.0, 168.0) |
| *Varying Passenger-Crew Contact Intensity* | | | | | |
| 50% Reduction | 849.0 (352.9, 1379.1) | 545.0 (230.0, 868.0) | 125.5 (54.0, 203.0) | 87.0 (31.0, 158.1) | 90.0 (31.0, 168.0) |
| 90% Reduction | 787.0 (332.9, 1346.1) | 535.5 (227.0, 899.0) | 96.0 (41.0, 173.0) | 62.0 (17.0, 130.0) | 87.0 (30.0, 170.0) |
| 100% Reduction | 744.0 (325.0, 1274.1) | 519.5 (225.9, 865.0) | 86.0 (37.0, 152.0) | 55.0 (17.0, 117.0) | 84.0 (29.0, 167.0) |
| *Varying Crew-Crew Contact Intensity* | | | | | |
| 50% Reduction | 897.0 (379.9, 1471.2) | 542.0 (220.8, 904.0) | 161.0 (70.0, 254.0) | 120.0 (48.0, 203.1) | 74.0 (23.0, 142.0) |
| 90% Reduction | 899.0 (404.0, 1529.2) | 558.0 (255.0, 943.2) | 165.0 (78.0, 274.0) | 118.0 (47.0, 206.0) | 61.0 (17.0, 132.0) |
| 100% Reduction | 895.5 (362.9, 1459.1) | 558.0 (218.0, 909.1) | 162.0 (68.0, 263.0) | 115.0 (44.0, 200.0) | 55.0 (15.0, 119.0) |
| **With Contact Intensity Reductions and PPE at Day 1, and No Network Lockdown** | | | | | |
| *Base Scenario* | | | | | |
| No Intensity Reduction | 3222.0 (2839.9, 3385.0) | 2311.0 (2091.0, 2402.0) | 472.0 (387.9, 526.0) | 151.0 (116.0, 191.0) | 280.0 (205.0, 335.0) |
| *Varying Passenger-Passenger Contact Intensity* | | | | | |
| 50% Reduction | 1623.5 (783.9, 2241.0) | 1012.0 (489.9, 1389.0) | 232.5 (110.0, 322.0) | 186.5 (85.0, 273.0) | 194.0 (81.0, 288.0) |
| 90% Reduction | 206.0 (44.0, 471.0) | 38.0 (9.0, 85.0) | 35.0 (8.0, 75.0) | 68.5 (12.0, 170.0) | 63.0 (11.0, 150.0) |
| 100% Reduction | 109.0 (14.0, 301.1) | 0.0 (0.0, 0.0) | 19.0 (3.0, 48.0) | 46.0 (4.0, 133.0) | 42.0 (3.0, 125.0) |

| | | | | | |
|---|---|---|---|---|---|
| *Varying Passenger-Crew Contact Intensity* | | | | | |
| 50% Reduction | 2925.0 (2454.8, 3144.1) | 2358.0 (2047.9, 2470.0) | 298.0 (216.0, 352.0) | 48.0 (30.0, 69.0) | 218.0 (131.0, 292.0) |
| 90% Reduction | 2526.0 (2092.9, 2709.0) | 2375.0 (1995.9, 2507.0) | 75.0 (46.0, 99.0) | 2.0 (0.0, 7.0) | 68.5 (23.0, 126.0) |
| 100% Reduction | 2375.5 (2042.9, 2498.0) | 2375.5 (2042.9, 2498.0) | 0.0 (0.0, 0.0) | 0.0 (0.0, 0.0) | 0.0 (0.0, 0.0) |
| *Varying Crew-Crew Contact Intensity* | | | | | |
| 50% Reduction | 3111.5 (2658.0, 3295.0) | 2319.0 (2059.8, 2414.0) | 518.0 (397.0, 584.0) | 131.0 (97.0, 166.0) | 139.0 (90.0, 178.0) |
| 90% Reduction | 3030.0 (2580.8, 3203.0) | 2331.0 (2040.8, 2423.0) | 555.0 (403.9, 632.0) | 116.0 (87.0, 148.0) | 26.0 (14.0, 40.0) |
| 100% Reduction | 3000.5 (2621.9, 3192.0) | 2324.0 (2077.8, 2428.1) | 558.5 (429.9, 650.0) | 114.0 (86.0, 143.0) | 0.0 (0.0, 0.0) |

**Supplemental Table 4.** Impact of Varying Intensity of Diagnosis-Based Case Isolation, with Asymptomatic Screening Starting at Day 15, Stratified by Network Lockdown and PPE Use, on COVID-19 Incidence and Mortality at 1 Month

| | **Cumulative Incidence** | | | **Cumulative Mortality** | | |
|---|---|---|---|---|---|---|
| **Scenario** | Total | NIA[1] | PIA[2] | Total | NDA[3] | PDA[4] |
| | *Median (95% SI)* | *Median (95% SI)* | *Median (95% SI)* | *Median (95% SI)* | *Median (95% SI)* | *Median (95% SI)* |
| *Network Lockdown and PPE at Day 15* | | | | | | |
| 100% Isolation | 928.0 (416.9, 1545.1) | 10.0 (-8.5, 30.5) | 1.1 (-0.9, 3.3) | 10.0 (3.0, 21.0) | 0.0 (0.0, 0.0) | 0.0 (0.0, 0.0) |
| 90% Isolation (Base) | 943.0 (411.8, 1556.0) | 0.0 (-20.5, 19.5) | 0.0 (-2.2, 2.1) | 10.0 (3.0, 21.0) | 0.0 (0.0, 0.0) | 0.0 (0.0, 0.0) |
| 75% Isolation | 958.5 (431.0, 1525.1) | -5.5 (-23.5, 14.0) | -0.6 (-2.6, 1.4) | 10.0 (3.0, 20.0) | 0.0 (0.0, 0.0) | 0.0 (0.0, 0.0) |
| 50% Isolation | 953.0 (414.9, 1548.0) | -16.5 (-35.5, 2.0) | -1.8 (-3.8, 0.2) | 10.0 (2.0, 21.0) | 0.0 (0.0, 0.0) | 0.0 (0.0, 0.0) |
| 25% Isolation | 974.5 (410.0, 1583.0) | -30.0 (-49.5, -9.0) | -3.2 (-5.3, -0.9) | 10.0 (3.0, 21.0) | 0.0 (0.0, 0.0) | 0.0 (0.0, 0.0) |
| No Isolation | 960.5 (426.9, 1588.0) | -28.0 (-45.5, -8.0) | -3.0 (-4.9, -0.9) | 10.0 (3.0, 21.0) | 0.0 (0.0, 0.0) | 0.0 (0.0, 0.0) |
| *No Network Lockdown or PPE* | | | | | | |
| 100% Isolation | 3499.0 (3251.0, 3591.0) | 18.0 (14.0, 22.0) | 0.5 (0.4, 0.6) | 32.0 (19.0, 46.0) | 0.0 (-1.0, 0.0) | 0.0 (-2.9, 0.0) |
| 90% Isolation (Base) | 3516.0 (3288.9, 3603.0) | – | – | 31.0 (19.0, 44.0) | – | – |
| 75% Isolation | 3544.0 (3373.0, 3617.0) | -27.0 (-30.5, -23.0) | -0.8 (-0.9, -0.7) | 32.0 (21.0, 46.0) | -1.0 (-1.0, 0.0) | -2.9 (-3.4, 0.0) |
| 50% Isolation | 3578.0 (3420.0, 3632.0) | -59.0 (-62.5, -56.0) | -1.7 (-1.8, -1.6) | 33.0 (20.0, 46.0) | -1.0 (-1.0, -1.0) | -3.2 (-3.9, -2.7) |
| 25% Isolation | 3601.0 (3492.9, 3644.0) | -81.0 (-84.0, -79.0) | -2.3 (-2.4, -2.2) | 32.0 (19.0, 46.0) | -1.0 (-2.0, -1.0) | -3.3 (-5.1, -2.9) |
| No Isolation | 3621.0 (3514.9, 3656.0) | -100.0 (-103.0, -97.0) | -2.8 (-2.9, -2.8) | 33.0 (20.0, 45.0) | -1.0 (-2.0, -1.0) | -3.3 (-5.3, -2.8) |

[1] Number of infections averted relative to base scenario

[2] Percent of infections averted relative to base scenario

[3] Number of COVID-related deaths averted relative to base scenario

[4] Percent of COVID-related deaths averted relative to base scenario

**Supplemental Table 5.** Impact of Timing of Mass Asymptomatic Screening and Diagnosis-Based Case Isolation, with No Network Lockdown and Stratified by PPE Use, on COVID-19 Incidence and Mortality at 1 Month

| Scenario | **Cumulative Incidence** | | | **Cumulative Mortality** | | |
|---|---|---|---|---|---|---|
| | Total | NIA[1] | PIA[2] | Total | NDA[3] | PDA[4] |
| | *Median (95% SI)* | *Median (95% SI)* | *Median (95% SI)* | *Median (95% SI)* | *Median (95% SI)* | *Median (95% SI)* |
| *Varying Timing of Mass Screening (Never PPE)* | | | | | | |
| Day 1 | 2286.0 (0.0, 3421.0) | 1403.5 (1396.0, 1409.0) | 38.0 (37.9, 38.1) | 7.0 (0.0, 24.0) | 29.0 (28.0, 29.0) | 81.2 (80.6, 81.8) |
| Day 5 | 2621.5 (16.0, 3353.1) | 1070.5 (1067.0, 1074.0) | 29.0 (28.9, 29.1) | 9.0 (0.0, 23.0) | 27.0 (27.0, 27.0) | 75.6 (75.0, 76.0) |
| Day 10 | 2917.0 (1787.8, 3310.1) | 775.0 (772.5, 777.5) | 21.0 (20.9, 21.1) | 13.0 (4.0, 25.0) | 23.0 (22.0, 23.0) | 63.6 (62.9, 64.1) |
| Day 15 | 2944.5 (2256.8, 3176.1) | 746.0 (744.0, 748.0) | 20.2 (20.2, 20.3) | 18.0 (8.0, 32.0) | 18.0 (17.0, 18.0) | 50.0 (48.6, 50.0) |
| Day 20 | 3102.5 (2588.8, 3360.1) | 590.0 (588.0, 591.5) | 16.0 (15.9, 16.0) | 30.0 (16.0, 45.0) | 6.0 (6.0, 7.0) | 17.1 (16.1, 18.4) |
| Day 25 | 3607.0 (3360.9, 3668.0) | 85.0 (84.0, 86.0) | 2.3 (2.3, 2.3) | 36.0 (24.0, 50.0) | 0.0 (-1.0, 0.0) | 0.0 (-2.5, 0.0) |
| Never (Reference) | 3692.0 (3679.0, 3699.0) | 0.0 (0.0, 0.0) | 0.0 (0.0, 0.0) | 36.0 (25.0, 49.0) | 0.0 (0.0, 0.0) | 0.0 (0.0, 0.0) |
| *Varying Timing of Mass Screening (Always PPE)* | | | | | | |
| Day 1 | 1629.5 (0.0, 3013.0) | 2012.0 (1998.0, 2023.0) | 55.3 (55.0, 55.4) | 5.0 (0.0, 20.0) | 27.0 (27.0, 28.0) | 85.2 (84.5, 85.7) |
| Day 5 | 1856.5 (12.0, 2837.4) | 1776.0 (1766.0, 1784.5) | 48.8 (48.6, 49.0) | 6.0 (0.0, 19.0) | 26.0 (26.0, 27.0) | 81.0 (80.5, 81.5) |
| Day 10 | 2240.5 (1058.0, 2815.1) | 1395.0 (1387.0, 1402.0) | 38.3 (38.2, 38.5) | 10.0 (2.0, 20.0) | 23.0 (23.0, 23.0) | 70.6 (70.0, 71.1) |
| Day 15 | 2372.0 (1585.6, 2755.0) | 1267.5 (1262.0, 1273.0) | 34.8 (34.7, 34.9) | 15.0 (5.0, 27.0) | 18.0 (17.0, 18.0) | 54.3 (53.5, 55.0) |
| Day 20 | 2656.0 (1980.9, 3033.0) | 983.5 (977.5, 988.5) | 27.0 (26.9, 27.2) | 26.0 (12.0, 40.0) | 7.0 (7.0, 8.0) | 22.2 (20.9, 23.3) |
| Day 25 | 3354.0 (2831.8, 3537.1) | 285.5 (282.0, 290.0) | 7.8 (7.8, 7.9) | 33.0 (20.0, 47.0) | 0.0 (0.0, 1.0) | 0.0 (0.0, 2.5) |
| Never (Reference) | 3643.0 (3563.0, 3669.0) | – | – | 33.0 (20.0, 45.0) | – | – |

[1] Number of infections averted relative to base scenario

[2] Percent of infections averted relative to base scenario

[3] Number of COVID-related deaths averted relative to base scenario

[4] Percent of COVID-related deaths averted relative to base scenario
